# Communicating risk of diabetes to patients results in lower incidence at 1- and 3-year follow-up and improved preventive intervention

**DOI:** 10.1101/2022.06.07.22274277

**Authors:** Lisa Bailey-Davis, G. Craig Wood, Carolyn F. McCabe, Adam Cook, David D. Rolston, Christopher Seiler, Christopher D. Still

## Abstract

**Objective:** One-third of American adults are estimated to have prediabetes but the vast majority are unaware. Accordingly, persons with prediabetes are equally unaware of their personal susceptibility of developing type 2 diabetes and the impact of weight loss on reducing the risk of progression. The primary aim of this study was to demonstrate that raising awareness of present risk, risk reduction, and offering available, accessible preventive interventions would result in fewer incidences of type 2 diabetes at 1- and 3-year follow-up periods. Secondary aims included clinician use of a novel diabetes risk calculator in primary care indicated by the proportion of patients being offered a preventive intervention.

**Methods:** In this single-arm study, persons with prediabetes from three primary care sites received a letter that communicated their personalized risk of progression to diabetes within three years; estimated risk reduction with 5, 10, 15% weight loss, reported in pounds, representing data illustrated in the diabetes risk calculator; and offered a choice of five free, 6-month, programs. Progression to type 2 diabetes was evaluated at 1- and 3-year follow-up periods using electronic health record data. Comparisons are made between the active participants and the non-respondent group. The diabetes risk calculator was implemented in 53 primary clinics in July 2020 and viewable in the electronic health record of persons with prediabetes without the need for clinician action. Electronic health record data were used to evaluate the rate of clinicians offering or referring persons with prediabetes to a preventive intervention—these rates of referral were compared between two phases (pre-implementation: 8/2019-2/2020; early implementation: 8/2020-2/2021).

**Results:** Persons with prediabetes (N=328) received letters and n=83 responded to the invitation to choose a preventive intervention (treatment). Follow-up data were available for n=55 treatment and n=169 non-respondents at 1- and 3-years. At 1-year follow-up, 2% of treatment and 20% of non-respondents developed type 2 diabetes (p<0.0001) and at 3-year follow-up, 25% of treatment and 42% of non-respondents developed diabetes (p=0.038). In the early implementation phase, N=8,771 persons had prediabetes, 0.4% received care, and 10.7% received a referral whereas in the pre-implementation phase, N=9,638 persons had prediabetes, 0.3% received care (p=0.088), and 9.7% were referred (p=0.029).

**Conclusion:** Progression to type 2 diabetes was halted at 1- and 3-year follow-up for a significantly higher proportion of persons who received diabetes risk calculator results and initiated a preventive intervention compared to those who did not. In the early implementation phase, clinicians referred a higher proportion of persons with prediabetes to preventive intervention when presented with diabetes risk calculator results as compared the pre-implementation period.

## Introduction

A bold public health objective has been established by Healthy People 2030 to reduce diabetes incidence from 6.5 to 5.6 per 1,000 adults but preventing diabetes among high-risk adults is challenged by strikingly low risk awareness.^1^ One in three US adults, 88 million, have prediabetes, yet nearly 90% are unaware that they have the condition.^2^ An estimated 33% of U.S. adults with prediabetes will develop type 2 diabetes in 5 years.^3-5^ Overweight and obesity are leading risk factors affecting 85% of persons developing diabetes.^6^ Consistent with Healthy People 2030, there are strategies to prevent or delay diabetes.

The landmark Diabetes Prevention Program (DPP) showed that intensive lifestyle intervention consisting of diet and physical activity modifications or metformin are both effective preventive interventions to prevent or delay diabetes.^7^ Relative to placebo, the reduction in persons developing diabetes was 58% for intensive lifestyle interventions versus 31% for metformin at 3-yr follow-up.^7^ Longer-term follow-up over a mean of 15 years among DPP participants found greater reduction for persons in the intensive lifestyle intervention than metformin groups (27% ILI vs 18% metformin) in diabetes incidence.^8^ The United States Preventive Services Task Force (USPSTF) identified a high strength of evidence from meta-analyses that intensive lifestyle interventions effected significant reductions in incident diabetes among persons who had overweight/obesity and prediabetes in trials ranging from 1 to 30-years of follow-up.^9^ Modest weight loss of 5-7% is clinically beneficial, but the risk of progression to type 2 diabetes is reduced with a greater magnitude of weight loss.^9,10^ A review of electronic health record (EHR) data for over 6,000 patients with obesity discovered that those with >15% versus <7% weight loss in a 2-yr index period had less than half the risk of developing type 2 diabetes at 10-yr follow-up.^11^

Effective preventive interventions like the DPP and medications are critical to prevent or delay type 2 diabetes, but awareness of prediabetes and the potential for risk reduction are prerequisites to activating a preventive intervention. We have previously described an algorithmic-based Diabetes Risk Calculator (Figure 1) to communicate a personalized baseline risk of developing diabetes in three years.^12^ Accordingly, we demonstrated that communicating the Diabetes Risk Calculator directly to patients, estimating risk reduction with weight loss, and offering a choice of free, effective preventive interventions resulted in an improved recruitment response rate, moderate participation rates, and improvements in weight and HbA1c levels at 1-year follow-up.^12^

**Figure 1.**
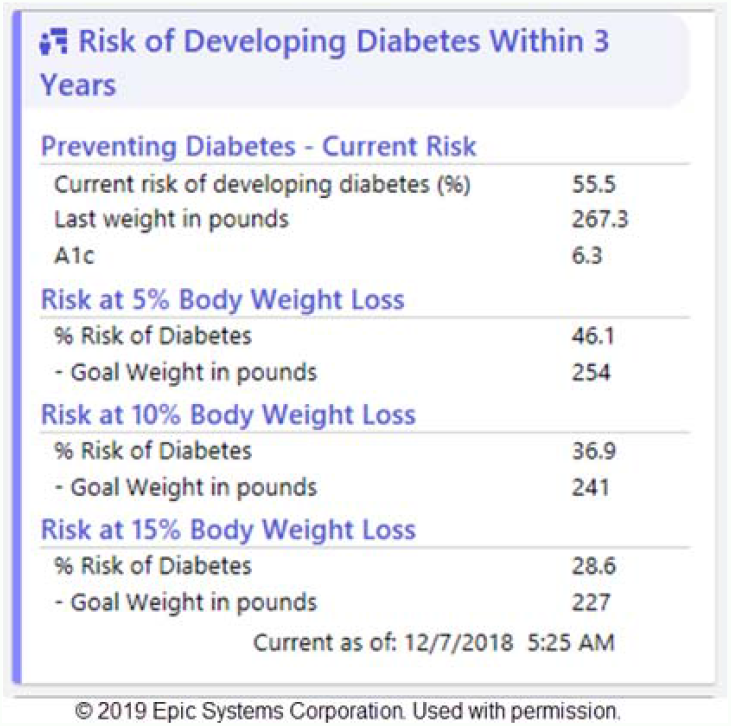
Diabetes Risk Calculator

In this paper, we report patient outcomes related to the incidence of type 2 diabetes at 3-year follow-up. Recognizing the need to improve patient awareness of diabetes risk, we also report early outcomes associated with implementing the Diabetes Risk Calculator directly into the EHR in primary care clinics. The USPSTF recommends screening for prediabetes and type 2 diabetes in persons aged 35 to 70 years who have overweight and obesity and clinicians should offer or refer persons with prediabetes to preventive intervetions.^13^ Given that the Diabetes Risk Calculator automates systematic screening, we evaluated the difference in the rate of care being offered or patients being referred to a preventive intervention from primary care clinics between the pre- to early-implementation phases.

## Methods

A single-arm pilot study was used to evaluate progression to type 2 diabetes at 3-year follow-up. All patients observed received care at Geisinger Health in central Pennsylvania during 2018-2022. Three primary care clinics were conveniently selected to participate in the pilot feasibility study to evaluate the impact of directly communicating the Diabetes Risk Calculator, risk reduction with magnitudes of weight loss (5%, 10%, 15%) in one year on diabetes risk at 3 years, and offering a choice of five preventive interventions including the Centers for Disease Control and Prevention (CDC)-approved DPP^14^ and medications. These methods have been fully described elsewhere.^12^ Eligible patients (N=328) were mailed recruitment materials. The response rate was 25.3% (n=81) and of these patients, n=62 qualified as having prediabetes at their baseline screening at study entry. These patients are considered the “treatment” group and are compared to patients in the “non-respondent” group (n=246) who received the recruitment materials but did not enroll in the study. The EHRs were evaluated at 3-year follow-up to determine incident diabetes among all patients. Indicators of diabetes included a problem list diagnosis of type 2 diabetes mellitus, anti-diabetes medications (excluding Metformin), and HbA1c value greater than or equal to 6.5%. Glucose levels were also evaluated with manual chart review to ensure that fasting measures were obtained; fasting glucose >126 mg/dL was considered an indicator for diabetes. In nearly all cases, multiple criteria were used to determine incident diabetes status. The percent that progressed to diabetes at 1- and 3-year follow-up periods was compared between the groups using Fisher’s exact test and a Type I error of 5%. Although 1-year outcomes were previously reported for the treatment group, a comparison was not made with the non-respondent group. Also, the treatment group evaluated herein had EHR data available at 1- and 3-year follow-up periods.

The impact of implementing the Diabetes Risk Calculator in primary care clinics (N=53) on the offering or referral of preventive interventions was evaluated using EHRs for patients who received care in two phases. The pre-implementation phase was August 2019 – February 2020. The Diabetes Risk Calculator was activated in the EHR in July 2020. The early implementation phase was August 2020 – February 2021. Clinicians received written communication about the Diabetes Risk Calculator prior to systemwide activation after the tool was briefly discussed at a service line virtual meeting. This was a less formal educational activity and notice than clinicians typically receive when a tool is introduced into care because system training experts were reassigned to systemwide COVID-19 mitigation efforts. The number of patients with ≥1 primary care encounter who met the criteria for the Diabetes Risk Calculator to fire (BMI >27, age ≥35 years, HbA1c 5.7-6.4%) were identified as the eligible population in each phase. Clinician offering of care was evaluated by prescription of anti-obesity medication (excluding Metformin). Clinician referral to a preventive intervention was evaluated for three service lines: Clinical Nutrition (Registered Dietitian/Nutritionist); Center for Nutrition and Weight Management (comprehensive behavioral, medical, and surgical interventions); and Wellness (CDC-approved DPP). Differences in preventive interventions being provided or prescribed between the pre- and early-implementation phases were evaluated using Chi-square test and a Type I error of p=0.05. Study procedures were reviewed and approved by the Geisinger Institutional Review Board (IRB# 2017-0394).

## Results

One- and 3-year follow-up data were available for n=55 patients in the treatment group and n=169 patients in the non-respondent group. There were no statistical differences between the groups in regard to age, sex, baseline BMI, hbA1c, or DRC (**Table 1**) Among those in the treatment group at 1-year follow-up, 11% had resolved prediabetes, 87% still had prediabetes, and 2% progressed to type 2 diabetes. At 3-year follow-up, 11% had resolved prediabetes, 64% still had prediabetes, and 25% progressed to type 2 diabetes. Compared to the non-respondent group, the treatment group had significantly lower rates of incident diabetes at 1-year (p<0.0001) and 3-year (p=0.038) follow-up. Among those in the non-respondent group at 1-year follow-up, 2% had resolved prediabetes, 79% still had prediabetes, and 20% progressed to type 2 diabetes. At 3-year follow-up, 5% had resolved prediabetes, 53% still had prediabetes, and 42% had type 2 diabetes (**Figure 2**).

**Table 1.** Baseline profile of study cohorts

|  | **Treatment**<br>**Participants**<br>**N=55** | **Non-**<br>**Respondents**<br>**N=169** | **Pre-**<br>**implementation**<br>**N=9,268** | **Early**<br>**implementation**<br>**N=8,771** |
| --- | --- | --- | --- | --- |
| Age, mean (SD) | 58.7 (9.4) | 60.1 (10.2) | 58.9 (14.0) | 58.8 (14.2) |
| Female, % (n) | 62% (n=34) | 49% (n=83) | 54% (n=5044) | 55% (n=4794) |
| BMI at baseline visit, mean (SD) | 35.8 (5.1) | 36.1 (6.3) | 37.6 (6.8) | 67.6 (6.7) |
| HbA1c at baseline visit, mean (SD) | 5.91 (0.17) | 5.92 (0.20) | 5.99 (0.21) | 5.99 (0.21) |
| Baseline DRC, mean (SD) | 15.5% (11.2) | 18.1% (15.8) | 23.7% (17.7) | 23.3% (17.5) |

\*significant difference between treatment and non-respondent groups
\*\* significant difference between pre-implementation and early implementation groups

**Figure 2.**
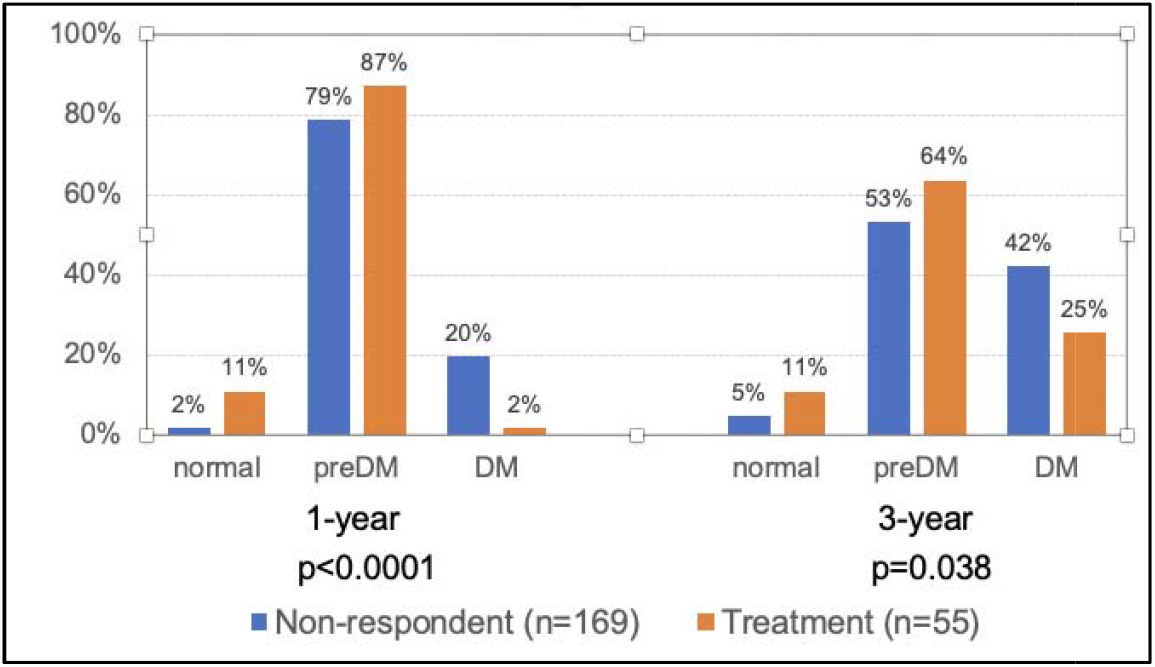
Proportion of persons with prediabetes at baseline who had resolved prediabetes (normal), maintained prediabetes (preDM), or had incident type 2 diabetes mellitus (DM) at 1- and 3-year follow-up periods

In the pre-implementation phase, N=9,268 patients met the criteria for the Diabetes Risk Calculator (not displayed in EHR). Of these, 0.3% received an anti-obesity medication and 9.7% received a referral to a preventive intervention; in total 9.9% received either intervention. In the early-implementation phase, there were N=8,771 patients who had the Diabetes Risk Calculator populate in their EHR during a visit. Of these, 0.4% received an anti-obesity medication and 10.7% received a referral to a preventive intervention; in total 11.0% received either intervention. Compared to the pre-implementation phase, a higher proportion of patients received a referral (p=0.029) and received either intervention (p=0.015) in the early-implementation phase (**Table 2**). Ultimately, systematic screening with the Diabetes Risk Calculator resulted in about 100 more persons with prediabetes being referred to a preventive intervention within the first 6 months of early implementation.

**Table 2.**
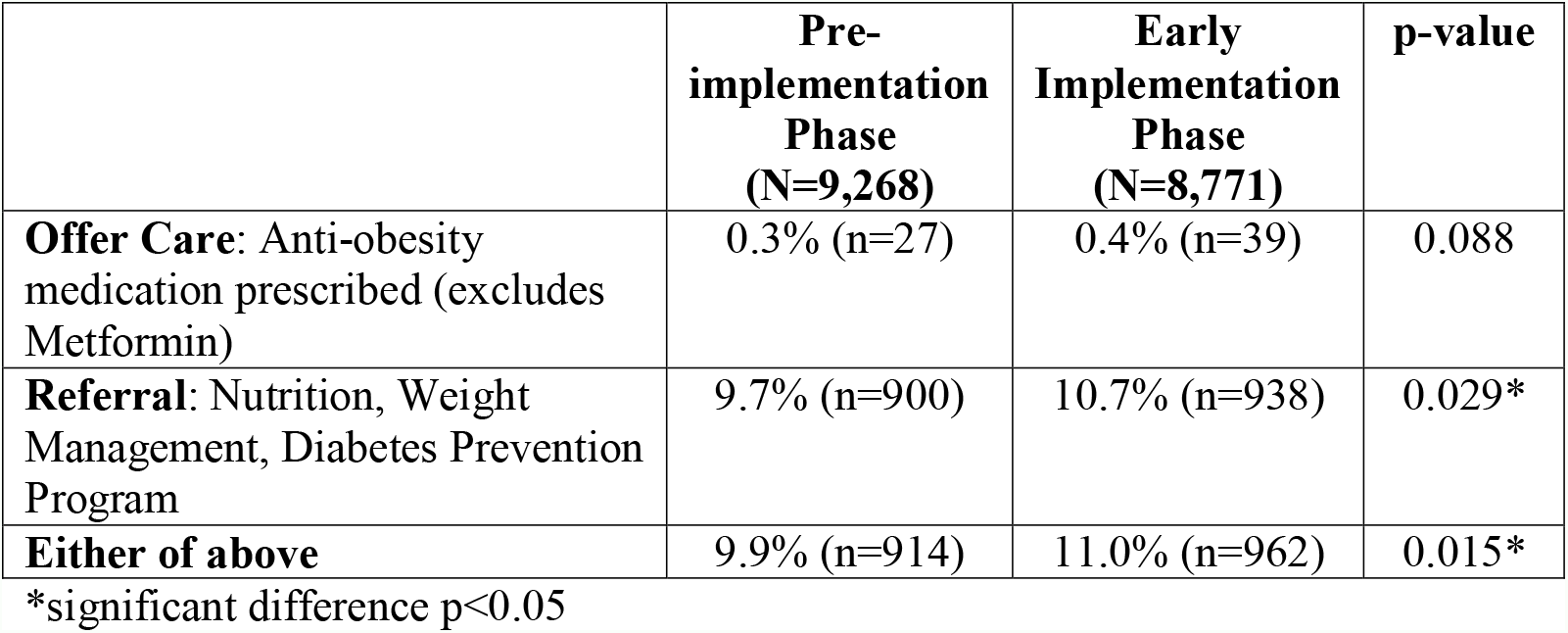
Comparison of patients being offered or referred preventive treatment prior to implementing systematic screening with the Diabetes Risk Calculator versus the early implementation phase

|  | **Pre-**<br>**implementation**<br>**Phase**<br>**(N=9,268)** | **Early**<br>**Implementation**<br>**Phase**<br>**(N=8,771)** | **p-value** |
| --- | --- | --- | --- |
| **Offer Care:** Anti-obesity medication prescribed (excludes Metformin) | 0.3% (n=27) | 0.4% (n=39) | 0.088 |
| **Referral:** Nutrition, Weight Management, Diabetes Prevention Program | 9.7% (n=900) | 10.7% (n=938) | 0.029* |
| **Either of above** | 9.9% (n=914) | 11.0% (n=962) | 0.015* |

\*significant difference $p<0.05$

## Discussion

Patients with prediabetes who received the Diabetes Risk Calculator results, a personalized baseline risk for developing type 2 diabetes with estimated risk reduction with weight loss, and acted on accessing a free preventive intervention of choice were significantly less likely to have prediabetes and diabetes at 1- and 3-year follow-up periods compared to patients who received the same information but did not access a preventive intervention. Further, when the Diabetes Risk Calculator was auto-populated in primary care, a significantly higher proportion of patients received USPSTF-recommended preventive intervention for prediabetes.^13^

These findings suggest that patients and clinicians who receive the Diabetes Risk Calculator are more likely to engage with or prescribe, respectively, with preventive interventions to delay diabetes. As such, the Diabetes Risk Calculator may advance efforts to achieve the Healthy People 2030 objective of reducing diabetes incidence.^1^ Scaling-up the Diabetes Risk Calculator to high-risk populations such as women with a history of gestational diabetes mellitus is a system-specific goal. Scaling-out the tool to other health systems is a strategy to remedy the observed barriers in screening. A systematic review found that prediabetes screening tools are not widely used due, in part, to reliance on patient or clinician entered data, questionable accuracy, and opportunity costs when screening interferes with patient-clinician interaction.^15^ Given that conceptual, structural, and cultural barriers, specifically weight stigma, interfere with clinicians’ communication of risk and prevention strategies, the Diabetes Risk Calculator appears to be an effective communication tool to increase referrals for obesity and diabetes prevention.^16-18^

## Data Availability

All data produced in the present work are contained in the manuscript

